# Multi-Platform Curation in the Development of ACMG/AMP Specifications for Von Hippel Lindau (VHL) Disease

**DOI:** 10.1101/2025.08.25.25334371

**Authors:** DI. Ritter, C Badduke, K Doonanco, HC Kang, T Pesaran, S Ridd, C Sheen, KM Farncombe, RH Giles, M Luo, N Pipko, CT Tsoi, K McGoldrick, C Mighton, RH Abu Kashabeh, MC Sanabria-Salas, Y Talab, KB Deka, MF Jacobs, E Tuzlali, B Gallinger, M Griffith, K Krysiak, J Machado, ER Maher, A Tirosh, RH Kim

## Abstract

**Introduction:** The Clinical Genome Resource (ClinGen) Von Hippel-Lindau (VHL) Variant Curation Expert Panel (VCEP) has created variant classification specifications tailored to the *VHL* gene, including phenotype-driven and evidence-based criteria, somatic and germline mutational hotspots, functional and in-silico data.

**Materials and Methods:** Using the American College of Medical Genetics and Genomics (ACMG) guidance and the ClinGen Sequence Variant Interpretation (SVI) recommendations, the VCEP made substantial modifications to eight evidence codes (PVS1, PS3, PS4, PM1, BS2, BS3, BS4, BP5), while 14 had minor or no changes and 6 were not used (PM3, PP2, BP1, PP4, PP5/BP6). The VHL VCEP applied two literature sets of over >428 papers in Clinical Interpretations of Variants in Cancer (CIViC) and >8700 structured annotations using Hypothesis.

**Results:** From 31 pilot variants, 15 remained pathogenic/likely pathogenic, 9 resolved to benign through the stand-alone benign evidence code and 7 variants with initial uncertain classifications, with many lacking additional literature, remained uncertain.

**Conclusion:** The versioned VHL VCEP specifications are publicly available in the ClinGen Criteria Specifications Registry and will enhance the transparency and consistency of variant classifications for this highly sequenced hereditary cancer gene.

## Introduction

Von Hippel-Lindau disease (VHL, OMIM 193300) is an autosomal-dominant hereditary cancer syndrome caused by germline pathogenic variants in the *VHL* tumor suppressor gene. Individuals with VHL disease have an increased risk of developing tumors and cysts in multiple organ systems, including clear cell renal cell carcinoma (ccRCC), hemangioblastomas in the retina and the central nervous system, adrenal pheochromocytoma and extra-adrenal paraganglioma, pancreatic neuroendocrine tumors, endolymphatic sac tumors, and cysts in the kidneys, pancreas, epididymis and broad ligament^1, 2^. While estimates vary, the syndrome has a prevalence of 1:39,000 to 1:91,111^3–6^ and a birth incidence of 1:35,000 to 1:42,987 worldwide ^3–6^

VHL is included on the American College of Medical Genetics and Genomics (ACMG) Secondary Findings list and some patients with a clinical VHL diagnosis may lack a *VHL* pathogenic variant, or conversely, may harbor a *VHL* variant without a VHL phenotype ^7^. Therefore, identifying germline pathogenic variants in *VHL* can inform clinical care^8^. For example, the risk of developing pheochromocytoma/paraganglioma (PPGL) is very low (VHL Type 1) among those with variants leading to an absent, unstable or truncated VHL protein; in contrast, there is a higher risk for developing pheochromocytoma (VHL Type 2) among those with a missense *VHL* variant affecting the surface of *pVHL*^9–11^. VHL Type 2 is further subdivided into families with a PPGL in addition to other VHL-related tumors (Type 2B) those with hemangioblastomas and PPGL but not RCC (Type 2A) and those with PPGL only ^11–13^. Of note, almost half of the single nucleotide *VHL* gene variants in ClinVar are classified as variants of uncertain significance (330/738 = 45%, with 99 conflicting interpretations, accessed 8/1/25), resulting in suboptimal genetic counseling and medical management.

To reduce uncertain and conflicting variant classifications., the Clinical Genome Resource (ClinGen) created the VHL Variant Curation Expert Panel (VCEP) to produce gene-based specifications to classify VHL variants. ClinGen is a large multi-centered effort through the National Genome Research Institute and National Cancer Institute that hosts a centralized resource of clinically relevant genomic information ^14^. ClinGen develops and supports expert panels to create variant classification specifications based on individual genes/diseases.^15^ In 2018, the ClinGen database was recognized by the Food and Drug Administration (FDA) as a Public Human Genetic Database, with formal training and documentation to evaluate variants within a versioned and transparent database (https://www.fda.gov/media/99200/download). To date, ClinGen has 37 approved VCEPs and over 10k variant classifications for 140 gene/disease specifications. Unique to the VHL VCEP are two collaborative efforts with online structured curation platforms - Hypothes.is and Clinical Interpretation of Variants in Cancer (CIViC) - which were used by the VCEP to facilitate variant classification ^16–19^. Here, we report on developing the VHL VCEP Specifications Version 1.1 and highlight the VCEP’s incorporation of structured variant data extracted from scientific literature across two platforms (FIGURE 1).

**Figure 1.**
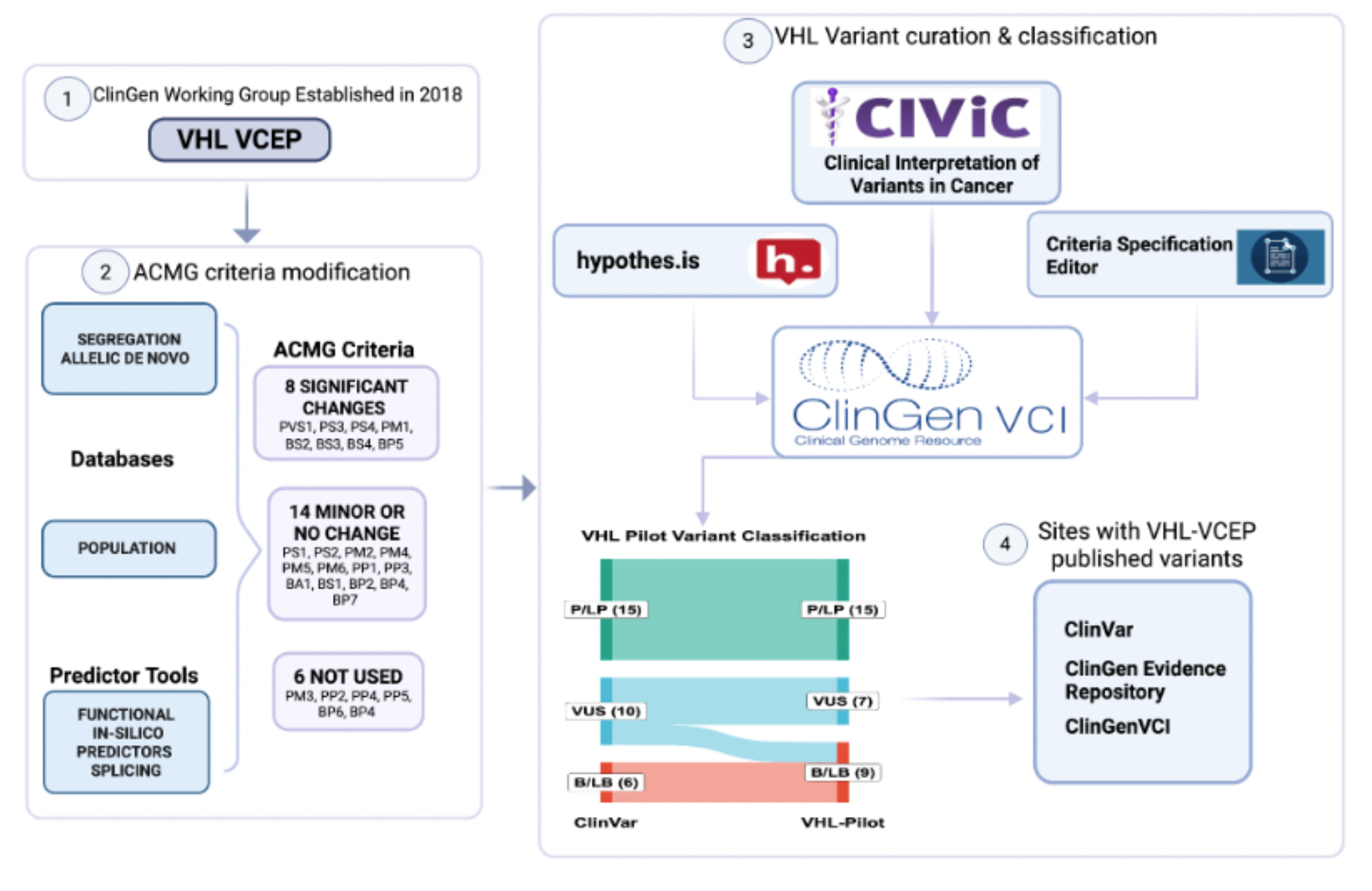
Overview of the ClinGen VHL VCEP Process. Step 1 indicates development and formation of the VHL VCEP. Step 2 highlights the sub-teams formed to work on specific sets of criteria and an outline of the evidence codes with significant or minor/no modifications as well as those not used. Step 3 shows the use of multiple curation platforms including the C-Spec Editor (3A), Hypothes.is and CIViC (3B) and variant curation in the ClinGen VCI, as well as the VHL VCEP Pilot results on 31 variants (3C). Step 4 depicts the shared data outlets: VHL Specifications in the C-Spec Registry (4A) and variants in the Evidence Repository and ClinVar (4B).

## Methods

### VCEP Formation and Organizational Overview

The VHL VCEP was established in 2018 to create VHL specifications and encourage sharing of clinical variant information^20^. The VCEP members span multiple clinical and research disciplines. VHL VCEP membership can be viewed on clinicalgenome.org: https://clinicalgenome.org/affiliation/50036. The VCEP is divided into three sub-teams with one focused on computational, functional and splicing, a second on segregation, allelic and de novo aspects and a third on population-related criteria (Figure 1, Sections 1-4). Draft specifications were entered in the ClinGen Criteria Specification Editor (CSpec Editor), a tool facilitating structured capture of variant classification specifications. The VCEP completed a pilot variant set (Step 3), followed by Step 4 approval and publication^21^.

#### In-Silico and Splice Predictors

112 pathogenic and benign clinically classified *VHL* variants were compared across in-silico predictors REVEL, SIFT and PolyPhen, and confusion matrices were used to evaluate potential thresholds. **Splicing analysis**: MaxEntScan, Splice AI and VarSeak scored 11 canonical pathogenic splice variants, 2 silent pathogenic splice variants and 1 clinically classified benign/likely benign intronic splice variant.

### Functional Domain Odds Analysis Assessing Evidence Code Strength

The VHL beta domain (63-155) containing the Nuclear Export domain (114-155), and the alpha domain (156-192) including the elongin C binding domain (157-172) cover much of the 3 *VHL* exons. For each domain in VHL, all variants, coding variants only, or missense variants only were extracted from ClinVar, and a simple odds calculation determined the proportion of benign/likely benign vs pathogenic/likely pathogenic in each domain.

### Pilot Variants, Biocuration and VCEP Approval Methods

#### Pilot Variants

The VCEP biocurators identified ~60 potential pilot variants from ClinVar^22^. We prioritized the following: Hypothes.is annotation count or CIViC evidence items, ClinVar submissions, and conflicting or uncertain (VUS) variant classifications. 31 *VHL* pilot variants were chosen including: 1 duplication, 9 frameshift and/or nonsense variants, 15 missense, 3 silent and 3 splice variants. **Biocurator Process:** Biocurators followed ClinGen Biocurator Training, documented in the ClinGen VCEP Protocol found on clinicalgenome.org^23^. The VHL VCEP used a two-curator review for 18 variants and single curator for 13 variants, followed by experts review with 22 responses captured through a Google form.

### Data Access

VHL Specifications are on the ClinGen Criteria Specification (C-Spec) Registry https://cspec.genome.network/cspec/ui/svi/affiliation/50036. All VHL VCEP published variants are on: (1) The ClinGen Evidence Repository, an FDA-recognized database of ClinGen variants https://erepo.clinicalgenome.org/evrepo/. (2) ClinVar, the NCBI database of clinical variants https://www.ncbi.nlm.nih.gov/clinvar/submitters/509654/ and (3) The ClinGen Variant Curation Interface (VCI), a public variant curation platform for germline variants https://curation.clinicalgenome.org/.

## Results

### VHL VCEP Specifications Overview

As VHL is an autosomal dominant condition with known pathogenic missense variants, the following evidence codes are not applicable: PM3, PP2, BP1; Per ClinGen, we do not use PP5 or BP6^24^. Additionally, the VHL VCEP uses PS4 case data instead of PP4. Substantial specification changes were made to the following: PVS1, PS3, PS4, PM1, BS2, BS3, BS4, BP5. Minor VHL-specific or no changes were made to the following: PS1, PS2, PM2, PM4, PM5, PM6, PP1, PP3, BA1, BS1, BP2, BP4, BP7. VHL-specific population thresholds for BA1, BS1 and PM2 were made following ClinGen SVI guidance or Ghosh et al ^25^. These are not discussed in detail but are found on the ClinGen C-Spec Registry.

### Evidence Codes Specified by the VHL VCEP

#### PVS1: Null variant (nonsense, frameshift, canonical +/−1 or 2 splice sites, initiation codon, single or multi-exon deletion) in a gene where loss of function (LOF) is a known mechanism of disease

The VHL VCEP has adapted the PVS1 decision tree following ClinGen SVI recommendations ^26^. PVS1 applies to truncating and splice-site variants at and after position 54. However, a loss of the first methionine in *VHL* (VHL p30) would not affect the second methionine at position 54 (VHL p19) and cannot be scored using the PVS1 decision tree. No other viable alternative starts are known. VHL protein critical domains include: the first beta domain (amino acids 63-155), which incorporates the nuclear export function (amino acids 114-155); the alpha domain (156-192), containing the elongin C binding region (157-172); and the second beta domain (193-204), crucial for protein stability (FIGURE 2). With only three relatively short exons, all *VHL* exon deletions receive PVS1. Loss-of-function variants after p.Met54 (positions 54-136) that lead to nonsense-mediated decay (NMD, after position 54 to the 5’ region of exon 2, position 138) or disrupt critical domains (positions 63-204) are classified as PVS1 ^27–29^. Truncations beyond the second beta domain (positions 205-213) resulting in minimal functional loss are categorized as PVS1_Moderate. Variants causing exon skipping or disrupting the reading frame between positions 63-204 are classified as PVS1, while PVS1_Strong is for severe disruptions (ex. reading frame disruptions) beyond position 205 or between positions 55-62, with the option to downgrade. Cryptic splice sites disrupting critical domains or predicted for NMD may receive PVS1, with PVS1 replacing PS3 for splice alterations.

**Figure 2.**
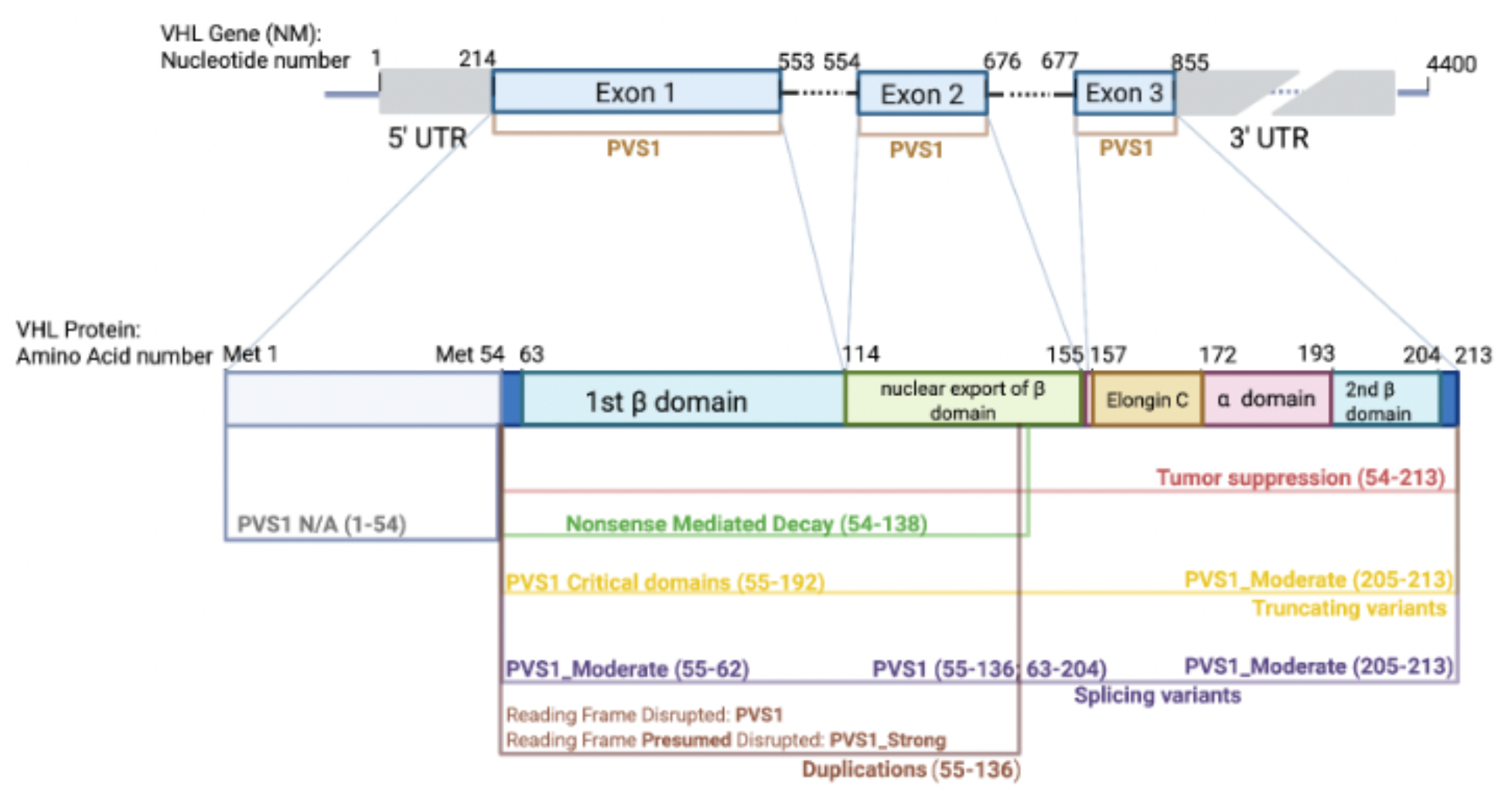
Major Functional Regions of the VHL gene are detailed and used to inform the full PVS1 decision tree found on the C-Spec Registry.

### Despite Large Scale Functional Data, Recommend Downgrading

#### PS3 to Supporting Strength PS3_Supporting

The VHL protein forms a complex with cullin 2 (CUL2), elongin B and C (ELB, ELC) and ring box protein 1 (RBX1), noted here as VCB-CR ^30^. The VCB-CR complex is active under specific cellular conditions and regulates the amount of hypoxia inducible factor HIF1a (and/or HIF2a, hereafter detailed in reference to HIF1a only). Under typical oxygen (normoxic) conditions, HIF1a is hydroxylated and bound by the VCB-CR complex, signaling ubiquitination and degradation. Under reduced oxygen (hypoxic) conditions, HIF1a is not hydroxylated, and the VCB-CR complex does not bind. Essential controls for HIF1a functional assays include: ensuring presence and activity of prolyl hydroxylases in normoxic conditions, VCB-CR formation, and proper functions of ubiquitination and proteasomal degradation (FIGURE 3). The VHL VCEP reviewed many small-scale experiments that lacked multiple controls and/or replicates (see guidance from Brnich et al through the ClinGen SVI ^31^). However, multiple studies with replication over time confirm the role of VHL in HIF1a regulation for VHL Type 1 and 2 A/B. The VHL VCEP suggests *VHL* functional evidence remain at PS3_Supporting for in-vitro assays that account for controlled cellular conditions and display total loss of HIF1a degradation (i.e. HIF1a presence). However, VHL Type 2C variants are typically missense variants in the alpha domain of VHL and do not usually affect HIF1a. Thus, if HIF1a presence is maintained and VHL Type 2C is suspected, assays evaluating fibronectin deposition or extracellular matrix assembly should be used.

**Figure 3.**
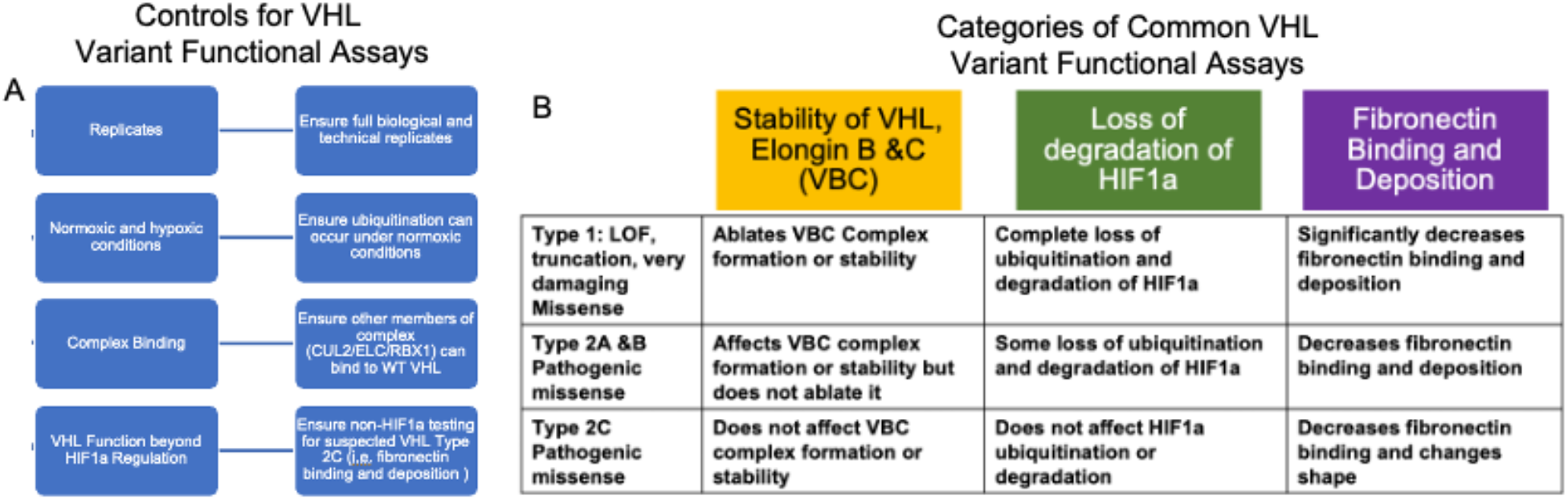
VHL Functional Assays and Controls. Figure 3A highlights some of the controls necessary for assays testing VHL variant function. Figure 3B shows common VHL functional assay types reviewed by the VCEP and their expected outcomes across VHL variant types. Examples of functional data reviewed include: (HIFa assays) Clifford et al 2001, Miller et al 2005, Hacker et al 2008, Rathmell et al 2004, Bangiyeva et al 2009, (protein stability) Rechsteiner et al 2001, and (fibronectin binding or deposition) Hoffman et al 2001.

A recent large-scale functional study of 2268 *VHL* variants applied a HIF1a assay using saturation genome editing (SGE) ^28^. We compared the results to 12 missense pilot variants: pathogenic (n=6), uncertain (n=4) and benign (n=2). As reflected by Buckley et al, the assay is best for loss of function or damaging missense variants (those associated with Type 1 VHL). Of 6 pathogenic missense variants, p.R64P was “intermediate” (−0.477) and p.R167Q scored substantially lower (−1.38 vs an average of −2.40 for the remaining 4 variants: p.S65W=−2.41, p.N78S=−2.35, p.P86L=−2.14, p.W88C=−2.68). Notably, for 5 /6 of the VCEP-classified pathogenic missense variants, additional functional data did impact the classification. For example, p.P86L lacked functional data in the curation and relied on multiple case data, but functional data does not increase a classification that is already at pathogenic. The 4 overlapping missense variants classified as uncertain by the VCEP ranged from neutral (2) to intermediate (1) and LOF2 (1). For 2 variants classified by the VCEP as benign using the Benign Stand-Alone criteria, the score was correspondingly low (−0.0025 and −0.053). While the authors note 95.2% sensitivity and 97.9% specificity discriminating between pathogenic and benign ClinVar variants, many of those variants include low-throughput HIF1a functional data contributing. To avoid circularity, the VHL VCEP aims to more fully evaluate this assay against variants classified without functional data.

### Use PS4 for Case Evidence instead of PP4, with a range from Supporting - Very Strong

#### PS4: The prevalence of the variant in affected individuals is significantly increased compared to the prevalence in controls

Given the rarity of VHL syndrome, the VHL VCEP uses PS4 to aggregate data from unrelated probands, and incorporates phenotype specificity (PP4, not used) by assigning lower points for “consistent but not highly specific” and “nonspecific” phenotypes. A diagnosis of VHL syndrome can be established through genetic confirmation of a pathogenic or likely pathogenic variant in the *VHL* gene or clinically, according to Danish, Dutch, or international criteria ^32, 33^. Probands meeting the Danish criteria receive 1 point (except for cases presenting solely with renal cell carcinoma (RCC) and pheochromocytoma, which are scored as “nonspecific” at 0.25 points if multi-gene panel is not performed. Phenotypes that are consistent with VHL but not highly specific (e.g., RCC and pheochromocytoma with an identified VHL variant) receive 0.5 points. Cases where the phenotype is not specified receive 0.25 points. The total points across the categories are added across unrelated probands to find PS4 strength (FIGURE 4).

**Figure 4.**
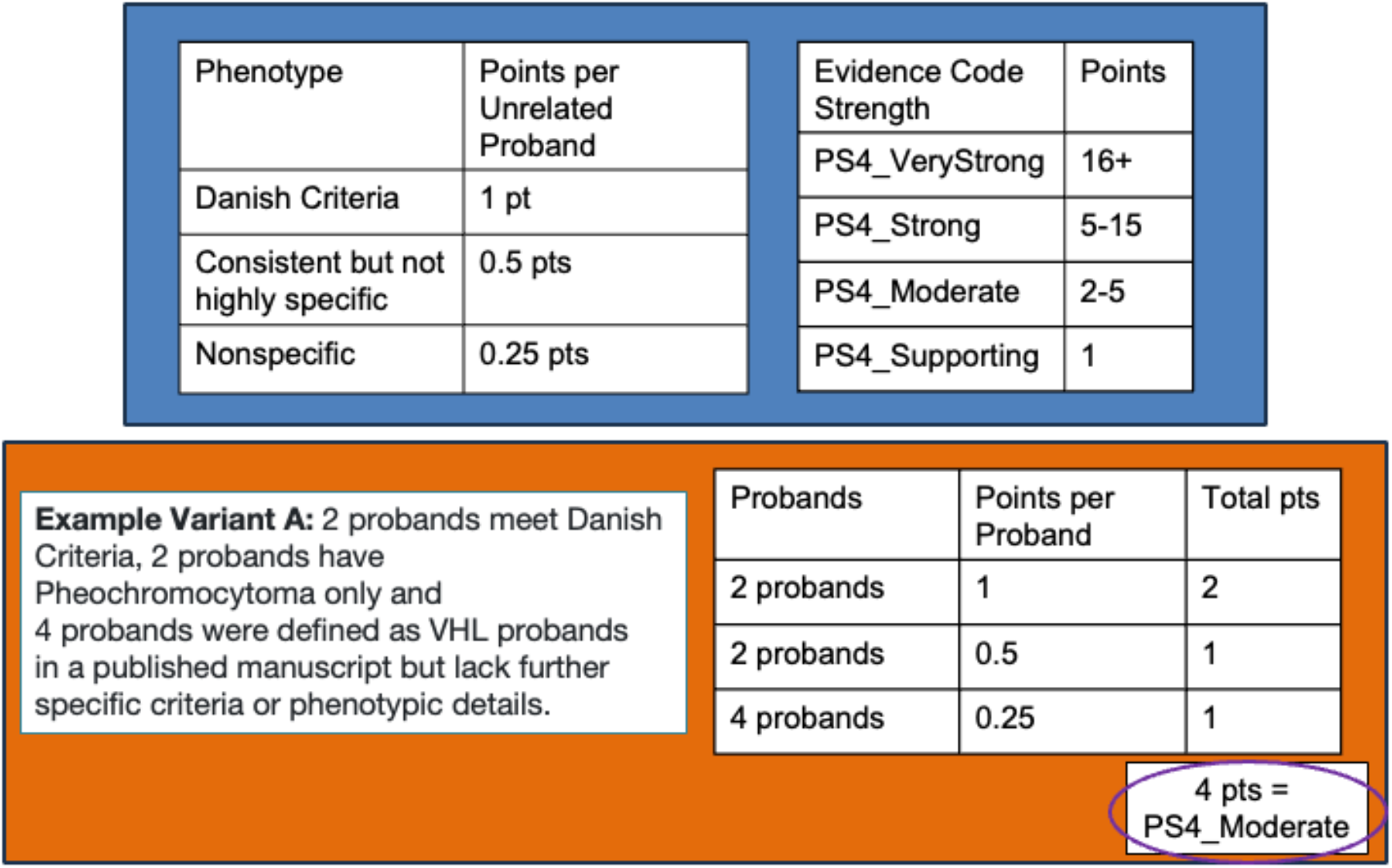
Combining phenotype and case-data into the PS4 Evidence Code. (Top Left) The VHL VCEP outlines points for highly specific, consistent but not highly specific and nonspecific cases, as well as (Top Right) additive strength across unrelated probands. (Bottom) An example of collected data for a variant and assigned additive points resulting in a PS4_Moderate strength for 8 total probands.

### Incorporating somatic cancer hotspots at supporting and moderate strengths

#### PM1: Variant in a mutational hot spot and/or critical and well-established functional domain (e.g. active site of an enzyme) without benign variation

PM1 can be used for known germline hotspots, and for 11 somatic hotspots (cancerhotspots.org) that do not overlap germline (see Walsh et al.) ^34^. The VCEP recommends following PM1 somatic hotspot strength where: amino acid (AA) positions with >= 10 somatic occurrences in the same AA receive PM1, while AA with <10 receive PM1_Supporting. Known germline hotspots, include those from Stebbins et al: R167, C162, L178, Y98, N78, P86, and population-specific hotspots such as E70K from Hwang et al as a hotspot in Korean cases (9/55 patients, 16.4%) and S65 (4.81%), R161 (5.35%), R167 (12.84%) from Hong et al in select Chinese cohorts^35–37^. Additionally, we use germline hotspots from the CIViC analysis of VHL literature reported by Chiorean et al, and available with VHL Specifications via the C-Spec Registry ^38^. The PM1 evidence code can also be applied to variants in critical functional domains if a residue is not a germline or somatic hotspot (see Figure 2 for critical functional domains). The VHL VCEP applied an odds calculation to variants in VHL functional domains and identified the following: first Beta Domain (156 (P/LP):15 (B/LB)= ~10.4, moderate) and Alpha Domain (66:7 = ~9.42, moderate), supporting use at moderate, though the second Beta Domain did not have enough variants for analysis.

### VHL Uses In Silico Computational Predictors with Supporting Strength

#### PP3: Multiple lines of computational evidence support a deleterious effect on the gene or gene product

ClinGen has identified REVEL, an ensemble-based meta-predictor, as a preferred tool for in silico missense predictions ^39^. An evaluation of REVEL, PolyPhen and SIFT using over 100 clinically classified VHL missense variants from ClinVar identified >=0.6 REVEL score as the best accuracy (ACC=0.926) with 91/92 P/LP and 22/30 B/LB correctly classified. PolyPhen and SIFT did not score above 0.80 accuracy, with PolyPhen (Hum-Var set only) performing better (ACC=0.79) than SIFT (ACC=0.55) on this small set. However, Pejaver et al, with the ClinGen SVI, analyzed REVEL scores and multiple in-silico predictors across most disease genes and identified a REVEL threshold of ≥0.664. ^40^ Although slightly more conservative, we adopted the Pejaver et al threshold while leaving in silico predictors at supporting. For splice variant in-silico evaluation, based on a small evaluation of performance on canonical and clinically classified splice variants, the VHL VCEP recommends using SpliceAI with scores ≥0.5 and VarSeak (class 4 or class 5) ^41^. Note that PP3 cannot be combined with PVS1, ensuring no redundancy in applying these evidence types.

### Benign Evidence Codes

#### Healthy Unaffected Adults over 65y with a VHL variant receive from Strong or Supporting Strength

#### BS2: Observed in a healthy adult individual with full penetrance expected at an early age

This criterion has been modified to reflect the lower penetrance of VHL at early ages, as full penetrance is not expected until age 65 ^5^. The strength depends on phenotyping and cancer screening for the absence of *VHL*-related tumors. BS2 can be applied when at least three unaffected individuals, all 65 years or older harbor the same variant and have undergone full phenotyping and cancer screening for the absence of VHL-related cancers. BS2_Supporting can be applied when full phenotyping and screening are unavailable. VHL disease-specific adaptation accounts for marked phenotypic variability and age-dependent penetrance ^42^.

#### BS3 Functional Data for benign variants

This evidence code can be used at full strength for functional tests of variants prior to position 54 that show the VHL p19 product is not impacted. Evidence of benign effect for VHL Type 1 and 2A/B can be evaluated when HIF1/2a (hereafter, referenced as only HIF1a) displays degradation (i.e. replicates WT function), and/or the VCB-CR complex stability is unaffected and/or extracellular matrix formation or fibronectin binding is unaffected. As VHL Type 2C variants typically do not affect HIF1a, absence of HIF1a alone for a suspected VHL Type 2C variant should not be used towards benign evidence. Functional studies displaying no impact on fibronectin or extracellular matrix formation are needed for VHL Type 2C. We recommend following the SVI guidance for functional assays, general and benign controls^31^. For splicing variants (and intronic/synonymous), RNA assays must demonstrate no impact on splicing.

A large-scale functional study using saturation genome editing and a HIF1a assay included benign and likely benign *VHL* variants from ClinVar ^28^. While the authors identify a threshold of >= −0.23 as “neutral”, about 7% of variants > −0.23 overlapped with known pathogenic variants, many of which were Type 2C. These variants are not candidates for HIF1a assays, as HIF1a presence is not reflective of the mechanism for VHL Type 2C. Due to the lack of true benign variants with clinical classifications made independently of small-scale functional assays in ClinVar, validating a benign functional score is challenging. However, we support use of the assay at supporting benign strength for suspected VHL Type 1 and Type 2 A/B variants, and not for Type 2C. The VHL VCEP plans to return to this data when more clinical benign variant classifications independent of functional assays are available.

### VHL Families lacking segregation can receive Strong or Supporting Strength

#### BS4: Lack of segregation in affected members of a family

For BS4 to be applied, affected members should meet the Danish Criteria for VHL^33^. Do not apply BS4 if family members are only affected with pheochromocytoma (e.g., Type 2C) and/or RCC. Families should be fully phenotyped for VHL manifestations to be considered unaffected. Given age-dependent penetrance, family members should also be at least 65 years old ^5^. If lack of segregation is observed in >=2 families, BS4 can be applied and BS4_Supporting for 1 family.

### Computational Predictors cannot be used for benign missense VHL variants, but Splice predictor can be applied at supporting strength

#### BP4: Multiple lines of computational evidence suggest no impact on gene or gene product

Due to the limited availability of benign variants and reduced classification accuracy for benign missense predictions, *missense predictors should not be used for benign supporting evidence in VHL*. Instead, BP4 can be applied only when computational evidence supports a lack of splicing impact. BP4 may be applied for variants with concordant splicing predictions indicating no impact. Specifically, SpliceAI scores of ≤0.1 and VarSeak classifications of Class 1 or Class 2 are required to qualify for BP4 ^41^.

### VHL variants with known alternative basis for disease can receive supporting evidence in clinical phenotypes do not overlap

#### BP5: Variant found in a case with an alternate molecular basis for disease

BP5 can be applied when a *VHL* variant co-occurs with a pathogenic variant from another gene that fully explains the phenotype, with conditions as follows: the alternative variant must be highly penetrant and consideration given to factors such as age, tumor type, sex, and family history (up to second-degree relatives). The proband and family clinical history should not overlap with VHL disease phenotypes. For example, BP5 would not apply to a patient with both a *VHL* variant and a pathogenic *SDHB* variant who has pheochromocytoma, as this tumor type is found in VHL disease. However, BP5 could apply in cases of chromophobe RCC where a pathogenic *FLCN* variant co-occurs, as VHL disease is not associated with non-clear cell RCC.

### VHL VCEP Pilot Variant Classifications Overview

Pilot variants were selected and curated by VHL biocurators (see Methods). Fifteen variants were initially classified as Pathogenic through at least two 1-Star laboratory classifications in ClinVar, and all 15 were confirmed as Pathogenic with the VCEP VHL specifications (Figure 2 Section 3). Six variants were initially classified as a range of Benign and Likely Benign and all were resolved to Benign, specifically reliant on the stand alone Benign (BA1) population frequency. Two variants had a mix of likely pathogenic and pathogenic classifications, and one was classified as Pathogenic (NM_000551.4(VHL):c.191G>C (p.Arg64Pro)) while the other remained Likely Pathogenic (NM_000551.4(VHL):c.264G>T (p.Trp88Cys)). Finally, for 10 variants where the initial classification was Uncertain by multiple 1-star laboratories, 7 VHL VCEP classifications were also Uncertain, with three moving to Benign. One variant, NM_000551.4(VHL):c.562C>G (p.Leu188Val), was classified as Uncertain although we noted it is likely a low-penetrance variant as follows “This variant has been reported as a putative low penetrance VHL type 2C variant; however at this time, the VHL VCEP does not have low-penetrance specific classification recommendations (see ClinGen Low Penetrance /Risk Allele Working Group)”.

### Use of Multiple Structured Biocuration Tools

The VHL VCEP is unique in that its biocurators contributed to two large sets of *VHL* scientific literature on independent structured curation platforms: CIViC (www.civicdb.org), with >630 *VHL* variants representing >428 papers, and Hypothesis (web.hypothes.is) with >8700 structured annotations from VHL clinical literature^16–19^. The dataset derived from CIViC’s open-access platform enabled analysis supporting VHL genotype-phenotype associations and identified candidate germline hotspot variants^38^. Similarly, Hypothes.is is an open-access web annotation tool that allows collaborative use, and ClinGen annotation protocols for Hypothes.is include highlighting defined variant and disease terms with structured text. A *VHL*-specific annotation protocol was established and published *VHL* cases were curated to supplement the VHL VCEP variant classification^15^. With Hypothes.is and CIViC as repositories of case-resolution VHL data, VHL VCEP operationalized the use of these two structured, curated literature sources into the specification development and pilot process. Variants were prioritized based on Hypothes.is annotation count, CIViC Evidence Items, the number of concordant pathogenic or benign ClinVar submissions, and/or conflicting/VUS ClinVar classifications. Our final pilot set (n=31) included 23 variants identified using Hypothes.is or CIViC curations and/or uncertain or conflicting ClinVar classifications. Notably, 29% (9/31) of these variants lacked curated literature in Hypothesis or CIViC. Among these, 55% (5/9) remained uncertain, compared to 9% (2/22) of variants with curated literature. This underscores the critical role of structured literature in variant classification. To facilitate integration of external curation resources into variant classification, the ClinGen VCI included a direct link to CIViC and Hypothes.is within the curation interface, through use of ClinGen Allele Registry IDs^43, 44^.

## Data Availability

All variants classified are available online at ClinVar or the ClinGen Evidence Repository.

https://erepo.clinicalgenome.org/evrepo/

## Discussion and Future Directions

The VHL VCEP has created the first version of specifications to aid VHL variant classification. These specifications, combined with use of the ClinGen VCI for structured evidence capture will increase the transparency and consistency of curated variants. The VCEP tailored several criteria to the VHL gene, including phenotypes for case-counting using PS4 and functional evidence, and functional data.

In the process, we applied two large sets of structured literature, created through the Hypothesis and CIViC platforms. The use of structured literature for variant classification emphasizes the need for scaled approaches, and careful prioritization, of literature curation. We found that a well-established pathogenic variant, NM_000551.4(VHL):c.583C>T(p.Gln195Ter), had extensive literature curations (>11 Hypothesis citations, >13 CIViC evidence items), when additional manuscripts will not alter the classification beyond pathogenic. Curation efforts for both platforms on the VHL datasets began approximately one year before the VHL VCEP developed a stable rough draft of specifications. Some criteria—such as BS2 with ≥3 individuals of >65 years— were not consistently annotated as the VCEP had not finalized the criteria. This highlights the importance of coordinating literature curation efforts with the VCEP process. We share the following considerations: (1) establish basic ACMG evidence code criteria before engaging in large-scale structured literature curation and (2) use preliminary variant assessments (from ClinVar submissions or non-literature evidence such as population or in-silico) to prioritize, reserving literature curation for variants where case or functional data may impact the clinical significance Overall, the parallel use of two structured, external literature curation sources was essential to our classification efforts - while offering key insights for future efforts integrating curated literature resources within ClinGen VCEPs.

ClinGen VCEP specifications are versioned and available in the ClinGen C-Spec Registry. Versioning assumes variant knowledge is additive – particularly regarding functional and clinical evidence – and the VHL Specifications will be updated over time. The VHL VCEP aims to thoroughly curate clinically relevant variants and re-assess large scale Saturation Genome Editing (SGE) functional assays ^28^. In addition, we note there are several reports of cryptic exons (E1) in VHL intron 1, and silent variants in exon 2 with evidence of exon skipping, and we plan to evaluate more silent and intronic variants ^45, 46^. There is also a need to consider somatic *VHL* variant classification informing cancer treatment for patients ^47^. We expect that the VCEP will iterate on these new specifications as we continue to improve the variant classification transparency and consistency for the *VHL* gene.

## Ethical Considerations

We follow the protocols of the ClinGen Data Access Privacy and Confidentiality WG (DAPC). All VCI users have attested to the data usage agreement within the VCI.

## Funding and Acknowledgements

Research reported in this publication was supported by the National Human Genome Research Institute of the National Institutes of Health under Award Number U24HG009649 (to Baylor College of Medicine /Stanford University). The content is solely the responsibility of the authors and does not necessarily represent the official views of the National Institutes of Health. In addition, this work is supported through the VHL Alliance and The Princess Margaret Cancer Foundation.

